# Effectiveness of a mHealth nutrition and lifestyle intervention (mHENAL) in reducing Cardiovascular Disease Risk in Sri Lankan adults: A Randomized Control Trial

**DOI:** 10.64898/2026.10.09.26365168

**Authors:** U Senarath, P Katulanda, NS Kalupahana, K Partheepan, M J Dibley

## Abstract

Cardiovascular diseases (CVD) are the leading cause of global mortality, responsible for 17.8 million deaths and 423 million prevalent cases annually. While the etiology of CVD is multifactorial, evidence confirms that multicomponent behavioral interventions regarding diet and lifestyle can substantially reduce risk. This study aimed to evaluate the effectiveness of a 12-month mHealth intervention, i.e., public health practice supported by mobile devices in reducing the 10-year CVD risk among overweight and obese adults aged 35 to 64 in an urban Sri Lankan population.

A two-group parallel-arm randomized control trial (RCT) was conducted with 1,200 participants from the Colombo district, assigned either to an intervention arm (n=600) or a usual care control arm (n=600). The intervention group received automated text and voice messages in their native language at a frequency of two of each per week over 12 months. The primary outcome, 10-year CVD risk, was assessed using the WHO CVD risk prediction score. The intervention significantly reduced the proportion of participants with a 10-year CVD risk from 10.0% to 6.2% (p=0.003), whereas no significant change was observed in the control group (11.0% to 9.7%). Participants in the mHealth arm demonstrated substantial behavioral improvements, including increasing daily fruit and vegetable servings from 3.6 to 5.5 and reducing daily salt intake from 7.64g to 5.62g. Physical activity levels also improved, with 89.6% meeting minimum daily requirements compared to 77.4% at baseline.

These findings suggest a potentially beneficial effect on predicted cardiovascular risk. The study concludes that an automated mHealth intervention is an effective, feasible, and acceptable approach to reducing cardiovascular risk in an urban South Asian setting. The findings provide a robust evidence base for integrating digital health strategies into national non-communicable disease (NCD) prevention policies. While the 12-month results are promising, further longitudinal research is required to evaluate the long-term sustainability of these behavioral changes and their ultimate impact on mortality rates.

**Competing interests:** There are no competing interests

**Author Approval:** All authors have seen and approved the manuscript.

## INTRODUCTION

Non-communicable diseases, particularly the cardiovascular diseases (CVDs) pose a major challenge to the sustainable human development in the twenty-first century. CVD is a broad group of diseases, predominantly consisted of ischaemic heart disease (IHD) and stroke. CVDs are the leading cause of death globally. According to the global burden of disease (GBD) estimates the total number of deaths due to CVDs was 19.8 million in 2022, which was approximately 32% of all deaths in the year (Mensah et al., 2023). IHD and stroke contributed to almost 85% of all CVD deaths, and 81.7% of total disability adjusted life years (DALY) due to CVD (Thomas et al., 2018). There is a rapid increase in morbidity and mortality due to cardiovascular diseases in Sri Lanka (Ministry of Health and Mass Media, 2024). The proportion of adults in the age 18-69 years with a history of a heart attack or chest pain from heart disease has doubled between 2015 and 2021, from 4.5% to 9.3% in men and from 4.2% to 8.5% in women. (Ministry of Health, Nutrition and Indigenous Medicine, 2016; Ministry of Health and Department of Census and Statistics, 2021).

The etiology of cardiovascular diseases is multifactorial and no single factor is an absolute cause (Smith, 2007). According to the INTERHEART study, abnormal lipids (OR=3.25), smoking (OR=2.87), hypertension (OR=1.91), diabetes (OR=2.37), abdominal obesity (OR=1.12), psychosocial factors (OR=2.67), consumption of fruits and vegetables (OR=0.7), alcohol (OR=0.91), and regular physical activity (OR=0.86) attributed to most of the CVD risk worldwide (Yusuf et al, 2004). Collectively, these nine risk factors accounted for 90% of the Population Attributable Risk in men and 94% in women. In Sri Lanka, diabetes, obesity and physical inactivity have been rising rapidly over the past 2 to 3 decades (Fernando et al, 2026). In particular the prevalence of overweight and obesity (BMI>25 kg/m^2^) has increased from 24.6% to 30.0% in adult men, and from 34.3% to 47.9% in adult women, between 2016 and 2021 (Ministry of Health 2016; 2021).

United Nations Sustainable Development Goals (SDG) target to reduce premature mortality from non-communicable diseases by one third between 2015 and 2030 (United Nations, 2015). Of the major NCDs, reduction in premature mortality due to CVD will gain the largest benefit in longevity in the population (Cao et al., 2018). It is assumed that multiple risk factor interventions using counselling and educational methods are efficacious and cost‐effective in reducing coronary heart disease (CHD) mortality and morbidity. A Cochrane review has concluded that interventions using counselling and education aimed at behaviour change do not reduce total or CHD mortality or clinical events in general populations, but may be effective in reducing mortality in high‐risk populations (Ebrahim et al., 2011). Another review highlighted that dietary advice is effective in bringing about modest beneficial changes in diet and cardiovascular risk factors, over approximately 12 months (Rees et al., 2013).

The unprecedented spread of mobile technologies as well as advancements in their innovative application to address health priorities has evolved into a new field of eHealth, known as mHealth. mHealth is defined as medical and public health practice supported by mobile devices, such as mobile phones, patient monitoring devices, personal digital assistants and other wireless devices (World Health Organization, 2011). With the evolution of mobile phone technology and its wider usage, mobile health (mHealth) is viewed as an attractive and promising approach to foster behavior change. There is a growing body of literature focused on mHealth interventions that address CVD risk factors. Despite the immense potential of mHealth interventions, evidence of their efficacy in mitigating cardiovascular risk is heterogeneous. Sri Lanka was aiming to achieve a 25% relative reduction in premature mortality from cardiovascular disease between 2015 and 2025 (Ministry of Health Nutrition and Indigenous Medicine, 2015). The present randomized controlled trial on mHealth was undertaken, considering the high burden of CVD, absence of individually focused nutrition and lifestyle counseling programmes, and the potential of use mobile phone technology in individually focused communications, The study aimed to investigate the effects of a mobile phone based nutrition counselling and lifestyle modification intervention (mHENAL) in reducing 10 year CVD risk among overweight or obese adults aged 35-64 years in the population.

## METHODS

### Study design

The study followed a stepwise approach for development and testing of mHealth interventions globally, which comprised of a: (i) Formative research; (ii) Compilation and pretesting the content; (iii) mHealth application development and pilot testing; (iv) Randomized control trial; and (v) Post-intervention evaluation. A two-group parallel-arm randomized control trial (RCT) was conducted. The RCT was a superiority trial with an allocation ratio of 1:1 for intervention and control groups, respectively. Scientific hypothesis of the study was whether mobile phone based nutrition counselling and lifestyle modification intervention (mHENAL) compared to usual care can reduce cardiovascular disease risk by one-third among individuals at increased cardiovascular risk (BMI ≥ 25 kg/m_2_, aged 35-64 years and living in a predominantly urban area) in a Randomized Controlled Trial.

### Participants

The study was conducted in Colombo district which has a population of 2,324,349, almost 11% of the country’s total population. It is the most urban district in Sri Lanka, with 78% of its residents living in urban areas (Department of Census and Statistics, 2012). Adults in the age 35-64 years, with a body mass index (BMI) of ≥25 kg/m_2_, and without any history of a cardiovascular event were included. Each participant was randomized either to intervention or control arm, using stratified block randomization method. Stratification was based on sex (female or male), age category (35-49 years or 50-64 years) and BMI category (overweight or obese). A sample of 1200 eligible participants from the Colombo district, who were assigned randomly either to mHealth (intervention arm, n=600) or usual care (control arm, n=600).

## Intervention

The intervention package contained a series of messages on nutrition and lifestyle practices aiming a change participants’ behaviour. These contents were prepared conforming to the best scientific evidence in principles and practice of nutrition and healthy lifestyles including recommended dietary modifications, food based dietary guidelines (Ministry of Health, 2011), portion control models (Jayawardena et al., 2017), and global recommendations on physical activity for health (World Health Organization, 2011a). The content and expression of messages were underpinned by relevant theories for behavior change applicable for m-health interventions (Cho et al., 2018).

The messages were focused on the following key themes: Healthy eating behaviour; calorie reduction; controlling dietary fats; sugar and salt restriction; increasing intake of fruits and vegetables; promoting physical activity; relaxation of mind and cessation of smoking and reducing alcohol intake.

The messages were pretested for clarity, usefulness, and relevance. The finalized messages were converted to both text and voice forms, recorded in male and female voices alternately in MP3 format, in Sinhalese and Tamil media separately. The text messages were formed in Unicode text, and were exactly the same as voice massages in Sinhalese and Tamil. The mHealth intervention encompassed two interconnected applications: a mobile application to register participants, and a web application to deliver text and voice messages. Automated voice and text messages were delivered to the mobile phone of the participants in the intervention arm, at a frequency of 2 text and 2 voice messages per week over 12 months. The participants in the control arm followed the usual care.

In order to operationalize message delivery, two interlinked applications, a mobile and a web application, were developed by a software development company (Data Management Systems Ltd.) in consultation with the investigators. Mobitel Private Ltd. was engaged as the mobile network provider to host and deliver the m-health intervention according to an automated schedule as per the web application. Figure 1 illustrates the communication pathways of the mHealth intervention.

**Figure 1.**
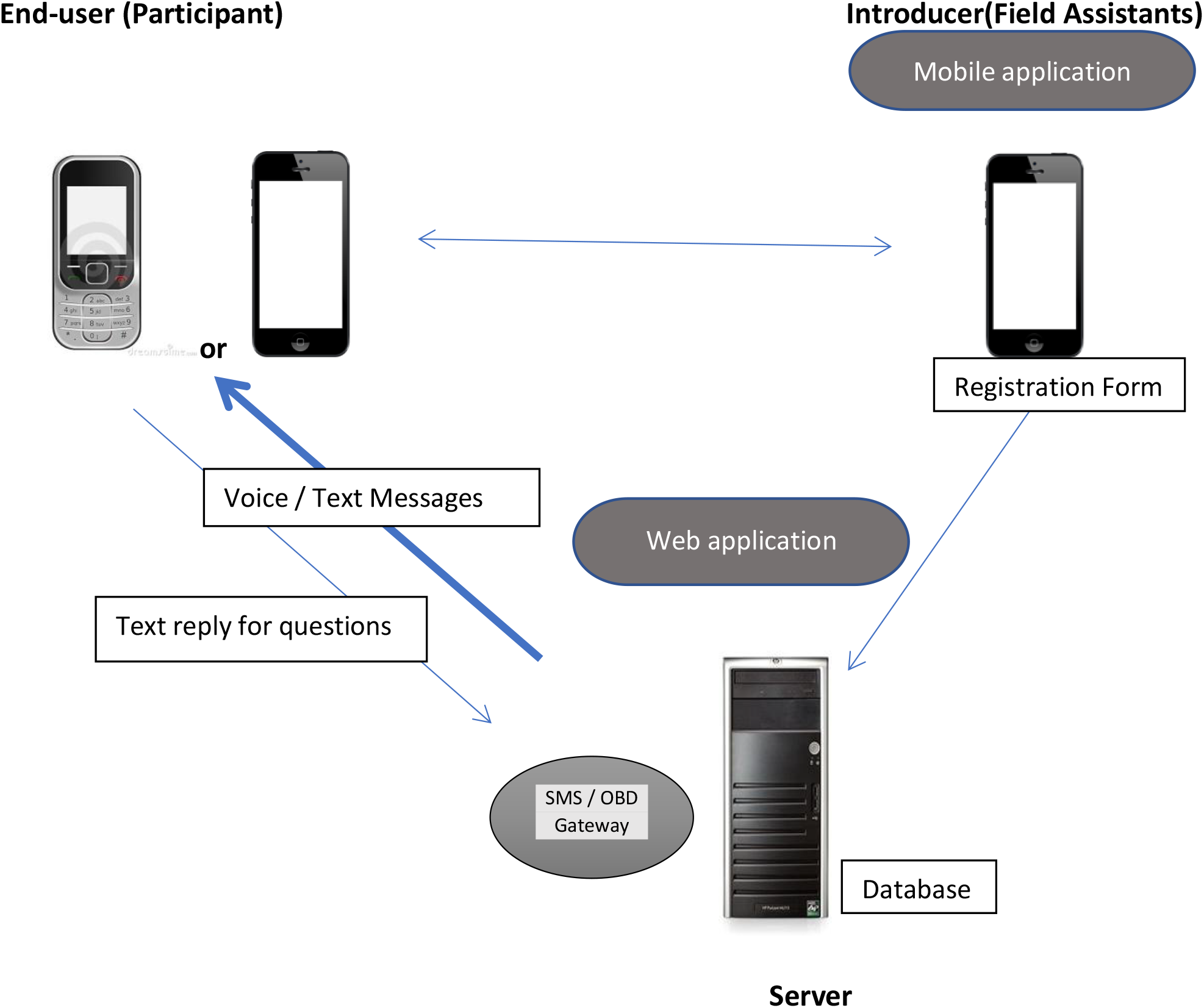
Flow diagram of the communication pathways of the mHENAL intervention

**Figure 2.**
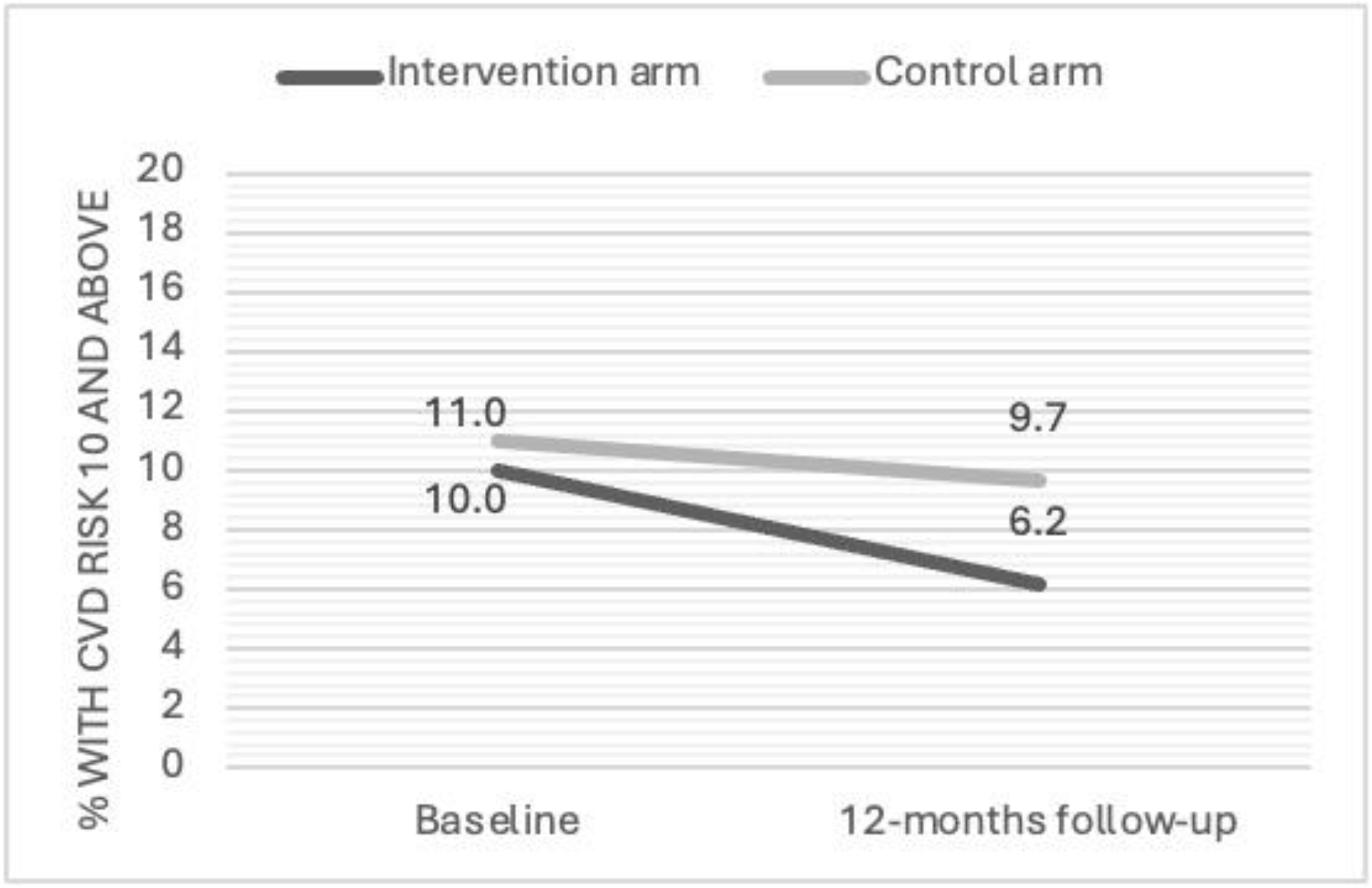
Percentage with WHO 10-year CVD risk of 10% and above in the intervention and control arms at the baseline and 12 months after mHealth intervention (n=964)

### Measurements

Data were collected at enrollment and after 12 months of the intervention on food consumption, physical activity, smoking, anthropometry, body composition, blood pressure, fasting plasma glucose, HbA1c and lipid profile. The primary outcome (10-year CVD risk) was assessed according to the recently validated WHO CVD risk prediction score using sex, age, smoking status, blood pressure, serum cholesterol and glycaemic status of the participants. Secondary outcomes were body weight, body mass index, body fat percentage, dietary intake and physical activity.

#### Questionnaire

A structured questionnaire was designed to obtain data on socio-demographics, smoking habits, alcohol consumption, chronic illness, dietary practices, dietary intake and physical activity. Dietary intake was assessed using food frequency questionnaire (FFQ) (Jayawardena et al., 2016). Physical activity was assessed by a short version of Physical Activity Questionnaire (Arambepola et al., 2007).

#### Anthropometry, body composition and blood pressure

Weight, height, waist circumference and hip circumference were measured by trained data collectors following standard procedure for anthropometric measurements. Digital weighing scale (Seca 813, Hamburg, Germany), stadiometer (Seca 213, Hamburg, Germany), and ergonomic circumference measuring tape (Seca 201, Hamburg, Germany) were used. Body composition was assessed by 4-point, bio impedance analyzer (Omron HBF 375, Kyoto, Japan). Blood pressure was checked by a medically qualified investigator using digital blood pressure apparatus (Omron HEM 7320, Kyoto, Japan).

#### Biochemical investigations

Blood samples (6 ml of blood per person) were drawn in 12-hour fasting state, and tested in 2 accredited laboratories using the recommended methods: HbA_1c_ using high-performance liquid chromatographic assay (Bio-Rad D10, USA); fasting plasma glucose using glucose oxidase method (Randox Imola, UK); total cholesterol using cholesterol oxidase method (Randox Imola, UK); HDL cholesterol by direct clearance method (Randox Imola, UK) and triglycerides by GPO-PAP method (Randox Imola, UK).

### Ethics and data safety

The study protocol was approved by the Ethics Review Committee of the Faculty of Medicine, University of Colombo (EC-16-061), and registered in the Sri Lanka Clinical Trials Registry (SLCTR/2016/018). A four-member data monitoring committee (DMC) comprising a medical specialist, biostatistician, monitoring and evaluation expert, and representative of an ethics review committee was established.

### Statistical analysis

Sample size was calculated using the primary outcome measure, the 10-year CVD risk using the formula described by Machin and Fayers (Machin & Fayers, 2011). Statistical analyses were performed using SPSS (V20.0). The primary outcome measure, the 10-year CVD risk, was calculated using updated WHO risk prediction charts (The WHO CVD Risk Chart Working Group, 2019). The secondary outcomes included intake of ‘healthy’ diet, physical activity, waist-hip ratio, body fat percentage etc. As per the intention-to-treat analysis, between-arm differences and within-arm individual-level changes were analysed using chi-square, Mc-Nemar, and t-tests. The analysis was extended to estimate the odds ratio for the effect on 10-year CVD risk (10-year CVD risk ≥10% vs <10%) by the intervention arm in contrast to the control arm. Regression analyses were performed using the endline 10-year CVD risk as the dependent variable, and the treatment arm (intervention or control) as the main predictor variable. Binary logistic regression (having 10-year CVD risk of ≥10% in contrast to <10%) analysis was performed to estimate the risk. The covariates included in the regression model were baseline CVD risk, sex, age category, marital status, education level, employment category, residential sector, body mass index, physical activity (METS adequacy) and ownership of a mobile phone.

The sub-group analyses by socio-demographic variables (sex, age, marital status, employment category, education level, and residential sector), current dietary practices, current physical activity level, pre-intervention BMI, and baseline status of having any NCD were carried out to examine whether effects are modified by such parameters. The analyses were extended to examine possible interactions too.

## RESULTS

### Baseline assessment of participants

As shown in Table 1, distribution of participants by sex, age category, ethnicity, level of education, type of employment, and residential sector were similar between intervention and control arms (p≥0.05). At the baseline, the proportions with individual CVD risk factors (smoking status, hypertension, diabetes, hypercholesterolaemia and low HDL levels) were not significantly different between the 2 arms (p≥0.05) (Table 2). The 10-year CVD risk was calculated according to the WHO CVD risk calculation chart recommended for Sri Lanka, using age, sex, smoking status, systolic blood pressure, diabetes status and total cholesterol or HDL levels. The 10-year CVD risk was and categorized as ≥ 10% and <10%. At the baseline, 10.0% participants in the mHENAL arm and 11.0% in the control arm reported a CVD risk ≥ 10%, (p≥0.05).

**Table 1.** Socio-demographic characteristics of trial participants by study arm (n = 1,200)

| Characteristic | Category | Intervention (n = 600), n (%) | Control (n = 600), n (%) | Total (n = 1,200), n (%) | p-value<sup>1</sup> |
| --- | --- | --- | --- | --- | --- |
| **Sex** | Female | 382 (63.7) | 383 (63.8) | 765 (63.8) | 0.952 |
|  | Male | 218 (36.3) | 217 (36.2) | 435 (36.2) |  |
| **Age group (years)** | 35–49 | 358 (59.7) | 357 (59.5) | 715 (59.6) | 0.953 |
|  | 50–64 | 242 (40.3) | 243 (40.5) | 485 (40.4) |  |
| **Ethnicity** | Sinhalese | 566 (94.3) | 580 (96.7) | 1,146 (95.5) | 0.227 |
|  | Tamil | 17 (2.8) | 8 (1.3) | 25 (2.1) |  |
|  | Moor | 16 (2.7) | 11 (1.8) | 27 (2.2) |  |
|  | Other | 1 (0.2) | 1 (0.2) | 2 (0.2) |  |
| **Education level** | None or primary (Grades 1–5) | 25 (4.2) | 29 (4.8) | 54 (4.5) | 0.183 |
|  | Secondary (Grades 6–11) | 162 (27.0) | 176 (29.3) | 338 (28.2) |  |
|  | GCE O/L passed | 128 (21.3) | 144 (24.0) | 272 (22.7) |  |
|  | GCE A/L passed | 180 (30.0) | 174 (29.0) | 354 (29.5) |  |
|  | Degree and above | 105 (17.5) | 77 (12.8) | 182 (15.2) |  |
| **Employment status** | Full-time public sector | 253 (42.2) | 238 (39.7) | 491 (40.9) | 0.483 |
|  | Full-time private sector | 74 (12.3) | 58 (9.7) | 132 (11.0) |  |
|  | Self-employed / own-account employer | 81 (13.5) | 95 (15.8) | 176 (14.7) |  |
|  | Casual employment | 5 (0.8) | 9 (1.5) | 14 (1.2) |  |
|  | Unpaid family work | 22 (3.7) | 20 (3.3) | 42 (3.5) |  |
|  | Retired | 8 (1.3) | 10 (1.7) | 18 (1.5) |  |
|  | Unemployed | 157 (26.2) | 170 (28.3) | 327 (27.2) |  |
| **Residential sector** | Urban | 421 (70.2) | 419 (69.8) | 840 (70.0) | 0.900 |
|  | Rural | 179 (29.8) | 181 (30.2) | 360 (30.0) |  |

<sup>1</sup> p-values are from chi-square tests comparing the distribution of each characteristic between the intervention and control arms. Values are presented as **n (%)**. GCE O/L = General Certificate of Education Ordinary Level; GCE A/L = General Certificate of Education Advanced Level.

**Table 2.** Comparison of cardiovascular risk factors in the intervention (mHealth) and control arms at baseline and endline.

| Parameter |  | Baseline |  | Endline |  |
| --- | --- | --- | --- | --- | --- |
|  |  | Intervention | Control | Intervention | Control |
|  |  | (n=600) | (n=600) | (n=500) | (n=465) |
|  |  | n (%) | n (%) | n (%) | n (%) |
| Smoking | No | 524 (87.3) | 531 (88.5) | 474 (94.8) | 438 (94.2) |
|  | Yes | 76 (12.7) | 69 (11.5) | 26 (5.2) | 27 (5.8) |
| Systolic blood pressure (mmHg) | <120 | 267 (44.5) | 279 (46.5) | 203 (40.6) | 156 (35.5) |
|  | 120-129 | 136 (22.7) | 140 (23.3) | 211 (42.2) | 193 (43.9) |
|  | 130-139 | 113 (18.8) | 84 (14.0) | 73 (14.6) | 72 (16.4) |
|  | 140-159 | 71 (11.8) | 75 (12.5) | 12 (2.4) | 19 (4.3) |
|  | ≥160 | 13 (2.2) | 22 (3.7) | 1 (0.2) | 0 (0.0) |
| Total cholesterol (mg/dl) | <160 | 66 (11.0) | 66 (11.0) | 146 (29.2) | 134 (28.8) |
|  | 160-199 | 202 (33.7) | 192 (32.0) | 174 (34.8) | 145 (31.2) |
|  | 200-239 | 211 (35.2) | 226 (37.7) | 117 (23.4) | 127 (27.3) |
|  | 240-279 | 97 (16.2) | 96 (16.0) | 51 (10.2) | 44 (9.5) |
|  | ≥280 | 24 (4.0) | 20 (3.3) | 12 (2.4) | 15 (3.2) |
| HDL (mg/dl)** | Normal | 327 (54.5) | 333 (55.5) | 309 (61.8) | 255 (54.8) |
|  | Low | 273 (45.5) | 267 (44.5) | 191 (38.2) | 210 (45.2) |
| Diabetes*** | No | 444 (74.0) | 420 (70.0) | 353 (70.6) | 325 (69.9) |
|  | Yes | 156 (26.0) | 180 (30.0) | 147 (29.4) | 140 (30.1) |

\* A person smoking any type of tobacco within past 30 days
\*\*HDL ≥ 50 mg /dl for women and HDL ≥ 40 for men was considered normal
\*\*\*Diabetes was defined as elevated levels of either fasting glucose (≥7.0 mmol/L [≥126 mg/dL]) or HbA1c (≥6.5%) or a person with a previous diagnosis of diabetes

### Effect of the intervention

Overall, the retention rate of the participants was 80.3% at the end of 12 months (964 out of 1200). Percentage of participants with individual risk factors that are used to calculate WHO CVD risk score (eg., smoking, systolic blood pressure, total cholesterol, HDL cholesterol, and diabetes status) is presented in Table 3. Marginal improvements were observed in the intervention (mHealth) arm, in contrast to the control arms.

**Table 3.** Ten-year CVD risk in the intervention and control arms at 12 months after mHealth intervention, stratified by socio-demographic characteristics and the baseline CVD risk.

| Variable | Subgroup | Intervention arm |  | Control arm |  | Chi-square & p-value |
| --- | --- | --- | --- | --- | --- | --- |
|  |  | n (%) | subtotal | n (%) | subtotal |  |
| **Sex** | Female | 7 (2.2) | 322 | 19 (6.2) | 306 | $\chi^2 = 6.437$, p=0.011 |
| | Male | 24 (13.5) | 178 | 26 (16.5) | 158 | $\chi^2 = 0.584$, p=0.445 |
| **Age group (years)** | 35–44 | 4 (2.1) | 189 | 1 (0.6) | 160 | $\chi^2 = 1.365$, p=0.243 |
| | 45–54 | 5 (2.3) | 215 | 10 (5.3) | 189 | $\chi^2 = 2.474$, p=0.116 |
| | 55–64 | 22 (22.9) | 96 | 34 (29.6) | 115 | $\chi^2 = 1.186$, p=0.276 |
| **Education level** | Below GCE O/L | 13 (8.4) | 155 | 24 (15.8) | 152 | $\chi^2 = 3.821$, p=0.051 |
| | GCE O/L passed | 5 (4.7) | 107 | 12 (11.5) | 104 | $\chi^2 = 3.238$, p=0.0720 |
| | GCE A/L and above | 13 (5.5) | 238 | 9 (4.3) | 208 | $\chi^2 = 0.315$, p=0.575 |
| **Residential sector** | Urban | 23 (6.5) | 354 | 31 (9.7) | 321 | $\chi^2 = 2.284$, p=0.131 |
| | Rural | 8 (5.5) | 146 | 14 (9.8) | 143 | $\chi^2 = 1.909$, p=0.167 |
| **Baseline CVD risk<sup>a</sup>** | Low | 10 (2.2) | 450 | 13 (3.1) | 413 | $\chi^2 = 0.725$, p=0.395 |
| | High | 21 (42.0) | 50 | 32 (62.7) | 51 | $\chi^2 = 4.346$, p=0.037 |
| **All participants<sup>b</sup>** | - | 31 (6.2) | 500 | 45 (9.7) | 464 | $\chi^2 = 4.056$, p=0.044 |
|  |  | OR=0.615<br>(95% CI 0.382-0.991) |  | OR=1.00<br>(reference) |  |  |

GCE O/L = General Certificate of Education Ordinary Level; GCE A/L = General Certificate of Education Advanced Level.
<sup>a</sup> WHO CVD risk at baseline was defined as “Low” if <10% and “High” if ≥10%.

At the end of 12-months, the proportion with high CVD risk reduced from 10.0% to 6.2% in the intervention arm (n=500, p=0.003 by McNemar test), and 11.0% to 9.7% in the control arm (n=464, p=0.377 by McNemar test) (Figure 4).

Ten-year CVD risk in the intervention and control arms within key socio-demographic strata are summarized in Table 3. At 12 months, 31 of 500 intervention participants (6.2%) and 45 of 464 control participants (9.7%) had a predicted 10-year CVD risk of at least 10%, corresponding to an unadjusted odds ratio (OR) of 0.615 (95% CI 0.382–0.991). The Odds Ratio for the effect of intervention, after adjusting for age, sex, education level and baseline CVD risk was 0.508 (95%CI:0.270, 0.955). (Table 4). Sub-group analyses indicated that individuals with high CVD risk at the baseline significantly improved their CVD risk following the intervention: substantial reductions were observed in males (from 20.8% to 13.5%, p=0.019), persons aged 55-64 years (from 36.5% to 22.9%. p=0.011), full-time private-sector employees (26.3% to 10.5%, p=0.022), and those who were self-aware of having NCD (15.9% to 7.7%, p=0.001).

**Table 4.** Effect of the mHealth intervention on 10-year cardiovascular disease risk: Final multivariable binary logistic regression model for endline results (n = 964)

| Variable | Category | Adjusted OR (95% CI) | *p*-value |
| --- | --- | --- | --- |
| Study arm | Intervention | **0.51 (0.27–0.96)** | **0.035** |
|  | Control | 1.00 (Reference) | — |
| WHO CVD risk at baseline | <10% | **0.06 (0.03–0.12)** | **<0.001** |
|  | ≥10% | 1.00 (Reference) | — |
| Sex | Female | **0.34 (0.18–0.64)** | **0.001** |
|  | Male | 1.00 (Reference) | — |
| Age group (years) | 35–44 | **0.14 (0.05–0.39)** | **<0.001** |
|  | 45–54 | **0.16 (0.08–0.32)** | **<0.001** |
|  | 55–64 | 1.00 (Reference) | — |
| Education level | Below GCE O/L | 1.83 (0.90–3.73) | 0.097 |
|  | GCE O/L passed | 1.10 (0.48–2.51) | 0.830 |
|  | GCE A/L and above | 1.00 (Reference) | — |

**Note:** OR = odds ratio; CI = confidence interval; CVD = cardiovascular disease; GCE O/L = General Certificate of Education Ordinary Level; GCE A/L = General Certificate of Education Advanced Level. Adjusted ORs are from the final multivariable binary logistic regression model. Reference categories are indicated as **Reference**. Bold indicates $p < 0.05$ .
Dependent variable: CVD risk ≥10%; the reference category for dependent variable is: CVD risk <10%.
<sup>1</sup>Reference categories for independent variables (OR=1)

The participants have increased the number of daily servings of fruits and vegetables, from 3.6 to 5.5 in the intervention arm (p<0.001), and from 3.6 to 4.1 in the control arm (p=0.001). The intervention was effective in reducing the average daily salt intake, a reduction from 7.64 to 5.62 grams per person (p<0.05) in the intervention arm, in contrast to 7.58 to 7.13 grams per person in the control arm (p =0.124).

Concerning physical activity, the persons who achieve MET-minutes ≥600 per day (minimum daily requirement) have increased from 77.4% to 89.6% in the intervention group (p<0.001), and from 76.9% to 80.0% (p=0.076) in the control group. The current smoking rate in men has decreased following the intervention (from 24.2% to 14.0%; p=0.001) as well as in the control group (from 22.2% to 14.6%; p=0.001).

In the intervention arm, there were significant reductions in the systolic blood pressure (mean, from 124 mmHg to 122 mmHg), and diastolic blood pressure (mean from 80 mmHg to 79 mmHg). In the control arm, both systolic (mean, from 124 mmHg to 126 mmHg) and diastolic (mean, from 80 mmHg to 81 mmHg) blood pressure have increased significantly. The proportion with a normal HDL level based on sex-specific cut-offs, has increased significantly in the intervention arm (from 54.5% to 61.8%, p=0.022) in contrast to the control arm (from 55.5% to 54.8%, p=0.745).

There was no significant differences in the body weight (72.6 kg vs 71.3 kg, p=0.066), BMI (28.84 kg/m_2_ vs. 28.73 kg/m_2_, p=0.654), the glycaemic status (HbA1c, 6.297 vs 6.400, p=0.288) and the total cholesterol normal level (34.6% vs 36.6%, p=0.738), between the intervention and control arms, respectively. Within the participants who received mHealth intervention, those who had better compliance with the intervention (66.8% read at least one text and listened to at least once voice message per week) gained larger benefits than those who had poor compliance. No adverse events were reported among participants during the study period.

Among intervention completers, 42.2% reported difficulties replying to messages, 77.0% missed some voice messages, and 35.8% reported difficulties maintaining initial behaviour changes beyond six months.

## DISCUSSION

The present study found that an mHealth intervention with automated text and voice messages supporting nutrition and healthy lifestyle changes over 12-months was effective in reducing 10-year CVD risk in overweight or obese adults in a predominantly urban setting in Sri Lanka. Intervention participants had significantly a lesser probability of having ‘10 year CVD risk ≥10%’ in contrast to those with usual care, irrespective of their age, sex, education level and baseline CVD risk. The mHealth intervention was also effective in improving dietary (fruit and vegetable intake and salt reduction) and lifestyle behaviour (physical activity and smoking status). The analysis was based on participants with available outcome data, and differential loss to follow-up introduces uncertainty about the estimated intervention effect.

The present study assessed outcomes 12 months following intervention, and this duration was consistent with many past studies. mHealth exposure duration in previous studies, ranged from 3 months to 24 months, mostly 6 or 12 months. The present study was among the few studies that assessed the overall CVD risk (Leu et al 2015).

This study was undertaken in the backdrop of Sri Lanka’s efforts to achieve the Sustainable Development Goal (SDG) target of reducing premature deaths due to noncommunicable diseases by one-third by 2030 (United Nations, 2015). A higher trend of NCDs is observed both in urban and rural settings (Katulanda et al 2025; Dissanayake et al 2025). This intervention package has a high potential to be integrated to the existing primary health care system in Sri Lanka. There have been initiatives by the state health services to enhance primary health care services in the recent past, with special emphasis on reducing the burden of NCD. Healthy Lifestyle Centers (HLC) were established at PHCs by the Ministry of Health in 2011 as a noncommunicable disease (NCD) screening service for adults aged 35 years or more, at primary health care institutions (Mallawaarachchi et al., 2016). The cluster care system with defined catchment area for each primary health care (PHC) facility aims for high coverage of detection and management of CVD risk in the population. Since the PHC facilities, i.e. the Primary Medical Units and Divisional Hospitals have defined catchment population, introducing mHealth intervention through PHC has the opportunity to achieve a high population coverage and to monitor its impact. The other potential platform is the General Practice where the family practitioners can enroll their clients into mHealth intervention. The most beneficial sup-groups, such as men, 55-64 year-olds, full-time private sector employees and those known to have NCD may be easily captured through the General Practice.

There were several limitations in the study. Lack of midline assessment after 6 months was a limitation. Some of the key measurements related to behaviours such as dietary behavior, physical activity and smoking status were based on self-reporting, thus subject to reporting bias. Behavioural outcomes were partly self-reported, and improvements were not consistent across all anthropometric and biochemical measures. Nevertheless, the trial provides an important foundation for further evaluation of mHENAL under routine implementation conditions There was a large number of individuals with diabetes, hypertension and dyslipidaemia in the sample. Even though medication adherence is an important predictor of CVD risk, the mHealth intervention did not include messages to motivate adherence to medication. There were some technical issues which were realized after launching the intervention.

The present study did not include a cost analysis, which would otherwise be important for the health sector to understand the cost implications if a decision is to be made regarding scaling up for the population.

Though, the study showed positive results in a RCT, feasibility and strategies for implementation of the mHealth in wider population is not adequately explored by this study. It is also important to review the long term impact. Based on the results and global evidence, we recommend that the present mHealth intervention package be scaled-up through the primary health care system in Sri Lanka, after necessary modifications. The participants of this study need to be followed up further for long-term impact. Challenges remained with participant engagement and maintaining behaviour change. These findings highlight the need to investigate how participants can be better supported to engage with the programme and translate health information into sustained behaviour change.

## Data Availability

All data produced in the present study are available upon reasonable request to the authors

## Acknowledgment

This study was primarily funded by the National Research Council, Sri Lanka (NRC-15-145). Additional support was received from the University Research Grants (AP/3/2/2017/SG/08), Medical Research Institute (MRI 11-2018), and Global Health Research Unit on Diabetes and Cardiovascular Disease in South Asians. Data Management Systems Ltd. developed the mHealth application, and Mobitel (Pvt) Ltd. implemented the intervention under the guidance of the research team.

